# A novel immersive virtual reality environment for the motor rehabilitation of stroke patients: A feasibility study

**DOI:** 10.1101/2022.03.30.22273051

**Authors:** Giulia Fregna, Nicola Schincaglia, Andrea Baroni, Sofia Straudi, Antonino Casile

## Abstract

We designed and implemented an immersive virtual reality environment for upper limb rehabilitation, which possesses several notable features. First, by exploiting modern computer graphics its can present a variety of scenarios that make the rehabilitation routines challenging yet enjoyable for patients, thus enhancing their adherence to the therapy. Second, immersion in a virtual 3D space allows the patients to execute tasks that are closely related to everyday gestures, thus enhancing the transfer of the acquired motor skills to real-life routines. Third, in addition to the VR environment, we also developed a client app running on a PC that allows to monitor in real-time and remotely the patients’ routines thus opening the door to telerehabilitation scenarios.

Here, we report the results of a feasibility study in a cohort of 16 stroke patients. All our patients showed a high degree of comfort in our immersive VR system and they reported very high scores of ownership and agency in embodiment and satisfaction questionnaires. Furthermore, and notably, we found that behavioral performances in our VR tasks correlated with the patients’ clinical scores (Fugl-Meyer scale) and they can thus be used to assess improvements during the rehabilitation program. While further studies are needed, our results clearly support the feasibility and effectiveness of VR-based motor rehabilitation processes.

**Significance statement:** Approximately 80% of stroke patients suffer from a hemiparesis of the contralateral upper limb. Motor rehabilitation has been proven to be of key importance to regain, partially or totally, the impaired motor skills. Rehabilitation techniques are based on the repetitive and intense execution of simple motor behaviors. As such they can become taxing and cumbersome for the patients. This often produces non-adherence issues with an obvious negative impact on motor recovery.

Here we describe a novel immersive virtual environment for upper limb motor rehabilitation and we report the results that we obtained in a cohort of 16 stroke patients. Our system was designed to turn rehabilitation routines into engaging games and to allow the remote monitoring of the patients’ exercises thus allowing telerehabilitation.

All our patients showed a high degree of comfort in our immersive VR system and they reported very high scores of ownership and agency in embodiment and satisfaction questionnaires. Furthermore, and notably, we found that behavioral performances in our VR tasks correlated with the patients’ clinical scores (Fugl-Meyer scale) and they can thus be used to assess improvements during the rehabilitation program.

## 1 Introduction

Stroke is the second most common cause of death worldwide (Donnan et al., 2008; Feigin et al., 2009) and one of the main causes of acquired adult disability (Bonita et al., 2004; Warlow et al., 2008; WHO, 2003). In most patients, the acute illness produces long-term consequences for them and their families (Langhorne et al., 2011). In particular, brain damage produced by the stroke results in sensory, motor, and cognitive impairments that reduce the patient’s quality of life and social participation (E. L. Miller et al., 2010). At the motor level, stroke causes deficits in one of the upper limbs in more than 80% of patients acutely and for more than 40% of them, chronically (Cramer et al., 1997). The sensorimotor recovery of the affected upper limb is a key goal of post-stroke rehabilitation, especially in consideration of its crucial impact on the patient’s independence and quality of life (Pollock et al., 2014). The period immediately following a stroke is critical for regaining, at least partially, motor skills and if specific rehabilitation programs do not take place there, patients frequently incur in long-term disabilities and reduced quality of life (Patel et al., 2006).

Neurorehabilitation aims at stimulating neuroplasticity after brain injury with the final goal of maximizing motor recovery (Sampaio-Baptista et al., 2018), and it is essential to regain, partially or totally, the impaired motor functions. It has been found that, to achieve best results, motor rehabilitation must be based on repetitive and intensive tasks (Sampaio-Baptista et al., 2018). Specifically, the execution of repetitive task training, executed in sessions repeated several times per week over a period spanning several weeks to several months or even years, has been proven to be instrumental to increase upper limb functions in stroke patients (Veerbeek et al., 2014). Furthermore, good rehabilitation outcomes seem to be strongly and positively associated with the patient’s motivation and engagement (Langhorne et al., 2011). However, due to its very repetitive nature, neurorehabilitation can quickly become cumbersome for the patients and thus produce severe adherence issues, which negatively affect the rehabilitation outcome (Paolucci et al., 2012). It is thus of outmost importance to develop enjoyable yet challenging training procedures.

Gamification procedures have been proposed to make the tasks more entertaining for the patients. However, such “games” are mostly based on simple tasks to be executed on a computer screen and thus partially disconnected from everyday gestures and movements. On the contrary, task-specific and context-specific trainings have been proven to be key-features for the transferring of the acquired motor skills to real life (Maier et al., 2019).

All the above issues have been recently further exacerbated by the COVID 19 pandemic that, on the one hand, resulted in a large number of Covid patients needing motor rehabilitation procedures and on the other hand created the need to move out rehabilitation procedures from the hospital to focus limited clinical resources on the treatment of severe cases.

To address these problems, we leveraged the power of modern computer graphics to design and implement an immersive virtual reality (henceforth VR) environment for upper limb rehabilitation (Figure 1). This system solves all the major problems outlined above. Firstly, by leveraging the intrinsic flexibility of VR-generated environments we can present a variety of scenarios and tasks to the patients and keep them interested and focused on their rehabilitation tasks. Secondly, having the patient immersed in a full 3D environment allows us to create tasks that are closely related to everyday activities (e.g. reaching for a glass of water) thus ensuring a transfer of the acquired motor skills to real life. Thirdly, modern VR head-mounted displays are light-weight and compact and they could be easily used at home by patients. Thus, although we are presently testing our system in a clinical setting, it is already fully compatible with telerehabilitation scenarios.

**Figure 1.**
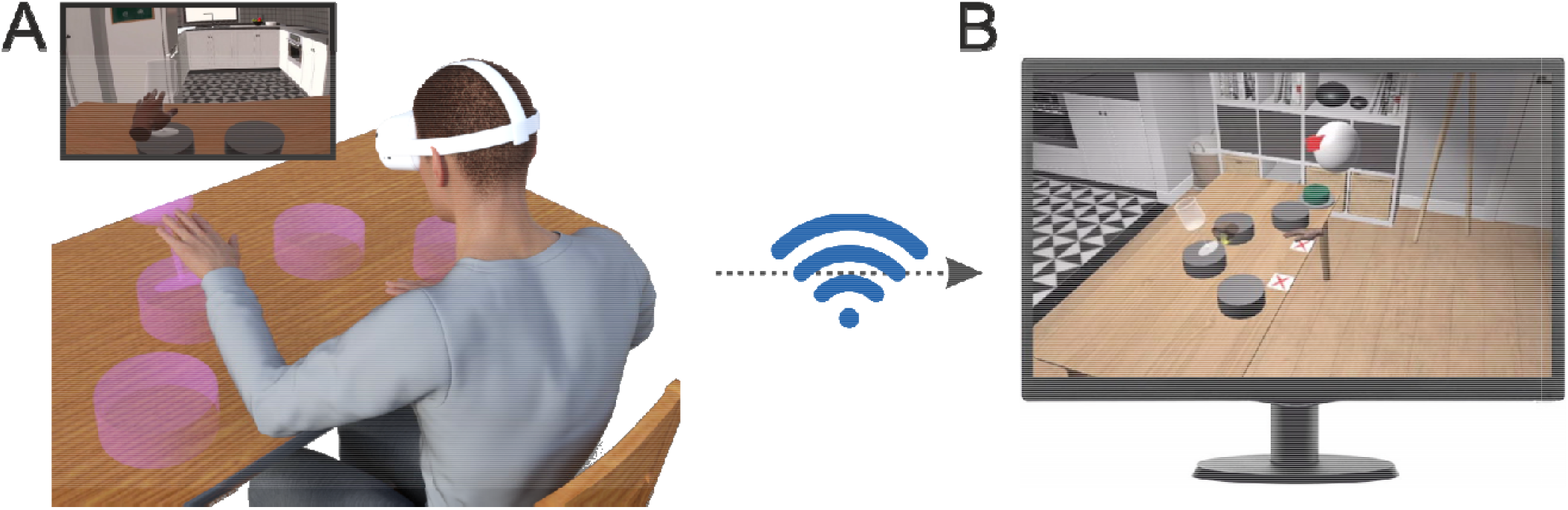
Application scenario of our immersive VR environment. (A) The patients a e immersed in a virtual environment by means of a head-mounted display (HMD, Oculus Quest 2, Facebook Reality Labs). In this environment, they can see different objects with which they can interact. The inset shows the scene as experienced by the patient on the HMD display. (B) The program running on the HMD wirelessly communicates with a client app running on a PC that allows to monitor remotely and in real-time the patients’ behavior, set their rehabilitation routin s and vocally interact with them.

Here, we describe the components of our system and report the results of a feasibility study in a cohort of 16 stroke patients. All our patients showed a high degree of comfort in our immersive VR system and they reported very high scores of ownership and agency in standardized embodiment questionnaires (Gonzalez-Franco & Peck, 2018). Furthermore, we found that behavioral performances in our VR tasks correlated with the patients’ clinical scores and they can thus be used to assess improvements during the rehabilitation program. We discuss these findings in the context of present and future clinical scenarios with an emphasis on telerehabilitation and on the potential combination of our VR environment with robotic devices presently used in rehabilitation procedures.

## 2 Materials and Methods

### 2.1 Subjects

16 subacute and chronic post-stroke patients (4 female, mean age 62±9) enrolled from the Rehabilitation Units of the Ferrara University Hospital participated in the experiments. They had a wide range of motor impairments and a diagnosis of first, ischemic or hemorrhagic stroke. No age restrictions were applied but patients affected by severe cognitive impairments or other co-existing clinical conditions were excluded. The clinical protocol and all procedures were approved by the local ethical committee (Comitato Etico di Area Vasta Emilia Centro (CE-AVEC) protocol code 897/2020/Oss/AOUFe approved on March 17^th^ 2021).

### 2.2 Experimental procedures

Prior to the experimental procedure, written informed consent was obtained from all patients. A clinical evaluation of the upper limb impairment and functioning was performed for all the included patients. All the assessments were conducted by the same trained physical therapist. The upper limb motor recovery was assessed by means of the Fugl-Meyer Assessment - Upper Extremity (FMA-UE) (Fugl-Meyer et al., 1975).

We also collected demographic and clinical information to characterize our cohort of patients with respect to age, sex, stroke type, hemiparesis side, days elapsed from the event and hospitalization type (i.e. inpatient or outpatient).

The results of clinical assessments and patients’ demographics are reported in Table S1 in the Supplementary Information.

### 2.3 Embodiment questionnaire

To evaluate the degree of embodiment of the virtual hands during the experiment we used a subset of a standardized questionnaire proposed by Gonzalez-Franco & Peck (2018). The questionnaire was administered in Italian at the end of the session and it consisted of 6 questions (see Supplementary Material). The patients could respond to each question by checking one out of 7 possible choices corresponding to a 7 point Likert scale ranging from -3 to 3, with -3 indicating strong disagreement and 3 indicating strong agreement with the statement Following Gonzalez-Franco & Peck (2018) we computed the Ownership and Agency indices by combining the questionnaire’s scores in the following manner:

1. Ownership: (Q1 - Q2) - Q3
2. Agency: Q4 + Q5 – Q6

### 2.4 Satisfaction questionnaire

At the end of each experimental session, we also administered a satisfaction questionnaire. The questionnaire was administered in Italian and it consisted of 10 items (see Supplementary Material). To six questions the patients had to respond by means of a 5-point Likert scale (1: not at all; 5: very much). Four questions had multiple-choices responses (see Supplementary Material).

### 2.5 Immersive virtual environment and client app

Our immersive virtual environment was developed in C# using the Unity 3D game engine (http://www.unity3d.com). It consists of two components: (1) A software package uploaded to the Quest 2 HMD that renders the virtual environment and manages the execution of the different tasks (Figure 1A) and (2) a client app running under Windows that wirelessly communicates with the HMD to manage the rehabilitation session (Figure 1B). Notably, the communication between the HMD and the client app takes place through the internet and thus these two devices can be, in principle, anywhere as they are not bound to be connected to the same local network.

As HMD we selected an Oculus Quest 2 for two main reasons. First, it has on-board capabilities that allow to optically track the patients’ hand movements. Second, it is lightweight and price-affordable.

The virtual environment consists of a cozy home interior (Scandinavian Interior Archviz purchased from the Unity Asset Store) selected to make the patients feel comfortable during task execution. In this environment, the patients use two virtual hands to execute different tasks with motor rehabilitation purposes.

During task execution, the client app provides the rehabilitation therapist with a real-time depiction of the virtual environment and the patients’ virtual hands as seen from a third-person point of view (Figure 1B). Furthermore, by means of a pop-up menu the therapist can in real-time and remotely manage the rehabilitation session by setting the tasks and the number of trials that the patient has to perform. The two virtual hands are controlled by the corresponding movements of the patients’ real hands. We also implemented a “mirror” condition in which one of the two virtual hands was controlled by the contralateral real hand. We performed preliminary tests of this condition in 6 patients that will not be reported here.

Four task are presently implemented in our system, which we called Glasses, Cloud, Ball in hole and Rolling Pin respectively. *Glasses*: The task starts with four pedestals presented on the table. The pedestals are distributed along a circle centered on the patient’s body at equal angular distances. A glass then appears on one randomly selected pedestal and the patients have to push it down (Figure 1A). The patients have to use the hand closer to the pedestal on which the glass appear (two pedestals are closer to the right hand and two are closer to the left hand). *Cloud*: At the beginning of trial a cloud of small bubbles, which pop upon touching, appears. The cloud is placed either to the right or to the left of the patients and they have to pop all of the bubbles with the corresponding hand. *Ball in hole*: For this task, a box-like support with a pocket at its center is placed on the virtual table. At the beginning of each trial a tennis ball is placed on this support either to the right or left of the patients and they have to gently push the ball into the hole with their corresponding hand. *Rolling Pin*: In this task, the patients have to use both hands to push for a pre-defined distance a rolling pin on the table. These four tasks were designed to re-create in the virtual environment rehabilitation routines usually performed in the clinical practice.

### 2.6 VR session

Upon coming to the lab, the patient was comfortably sit in a chair in front of a table. The experimenter then helped the patient to wear an HMD and immersed her/him in the virtual environment depicting a home interior. The patient was placed in front of a table also in the VR environment. The experimenter then used calibration routines programmed in our system to set the height and distance of the table in the VR environment to match those of the real table that the patient was facing. In this manner, when touching the table in the VR environment the patient also experienced a real sensation of touch produced by the real table. This step was implemented, based on previous results showing that the experience of multi-modal (in our case, vision, touch and proprioception) matching cues enhance the feelings of embodiment, presence and immersion of subjects in a VR environment (Gallace et al., 2012; Martin et al., 2022).

## 3 Results

Figure 1 graphically depicts the structure of our immersive VR motor rehabilitation system. The patients are immersed in a virtual environment containing different objects (Figure 1A) and their hand and finger movements, visually captured by the Oculus Quest onboard software, are used to animate two virtual hands through which they can interact with virtual objects (e.g. the magenta transparent glass in panel A) to perform different tasks (see Methods section for further details). During task execution, our system wirelessly communicates with a client app running on a PC that shows the scene in which the patient is immersed as well as her/his virtual hands from a third-person point of view (Figure 1B). Through this app the rehabilitation therapist can in real-time and remotely: (1) monitor the patients’ actions; (2) select the routine that they have to perform (e.g. task sequence, number of trials for each task, etc.); and (3) vocally interact with them. Notably, we designed our system such that the HMD and the client app need not be on the same local network, thus enabling telerehabilitation scenarios in which the patients can perform most of their routines at home while maintaining strict medical supervision.

Here, we report the results of a feasibility study that we performed in a cohort of 16 patients (4 female, mean age: 62±9, see Table 1). Each patient was tested once during the performance of multiple consecutive sessions, each consisting of four tasks (see Methods for a complete description of the tasks). The total duration of the VR-based training ranged between 40 minutes and 1 hour.

At the end of the experiment, all patients filled in a satisfaction and an embodiment questionnaires. In the satisfaction questionnaire, the patients had to rate in a scale from 1 (no satisfaction) to 5 (complete satisfaction) their subjective experience in the VR environment. Results in Figure 2A clearly show that almost all patients gave the maximum available score of 5 to their experience. The embodiment questionnaire evaluated the degree of ownership and agency produced by the virtual hands. Both scores range from a theoretical minimum of -9 to a maximum of +9 with positive values indicating increasing levels of embodiments. The average, across our patients, of the ownership and agency scores were both very close to their theoretical maximum (mean ownership= 7.4±2.0; mean agency= 8.3±2.0) and, of course, significantly different from 0 (ownership: p<<0.001, agency p<<0.001). In summary, the results of Figure 2 show that our immersive VR system for motor rehabilitation was highly appreciated by the patients and acting by means of virtual hands produced in all of them substantial subjective impressions of ownership of the virtual body and agency.

**Figure 2.**
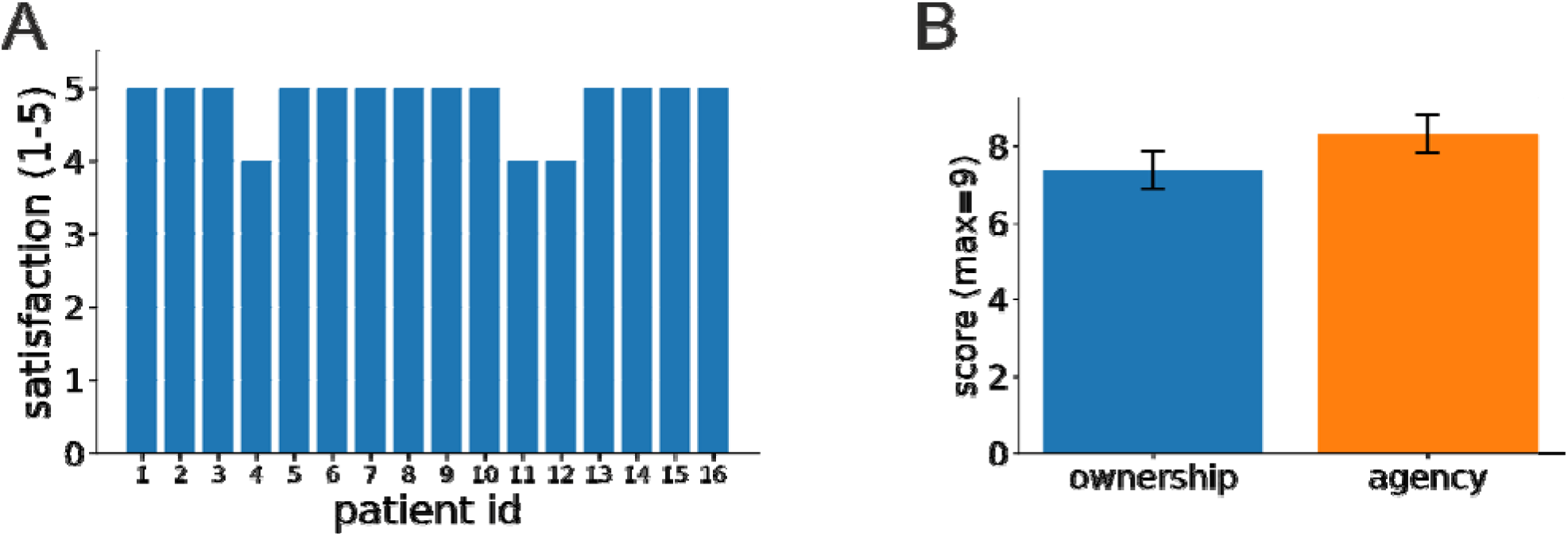
Patients’ feedback on their experience in our VR-based rehabilitation system. (A) Patients’ ratings, in a scale from 1 to 5, to the question: “Did you enjoy this type of training?” (In Italian: “Ha gradito la tipologia di allenamento?”). See Figure S2 in the Supplementary Material for the patients’ responses to the other points in the satisfaction questionnaire. (B) Patients’ scores for ownership and agency as assessed by a standardized questionnaire (see Methods for further details). The two bars represent average across patients and the vertical lines signify

A very promising use of our environment is that of automatically providing quantitative assessments of motor performance to the therapist to inform the rehabilitation process. This functionality is presently in an initial state and, due to continuous technical development of our system, was available only for a subset of 9 patients. It can nonetheless provide very useful information. To this end, Figure 3 shows the completion times for three tasks presently implemented in our system for two of our patients: a male patient in his 70’s (patient #12) and a female patient in her 60’s (patient #13). Completion times were computed from the onset of hand movement to task completion. As expected, in almost all conditions, completion times were significantly higher for the impaired compare to the healthy limb (patient 12: Ball in hole task: median left= 1.8s, median right = 2.17s, p<0.01; Cloud task: median left=3.45s, median right = 4.88s, p<<0.01; Glasses task: median condition 0 = 0.76s, median condition 1= 0.98s, median condition 2 = 1.12s, median condition 3 = 0.93s, p_0,3_=0.07, p_1,2_<0.01. Patient 13: Ball in hole task: median left = 2.91s, median right = 1.44s, p<<0.01; Cloud task: mean left=6.65s±0.88s, median right = 5.79s, p<<0.01; Glasses task: median condition 0 = 1.46s, median condition 1 = 2.15s, median condition 2 = 0.77s, median condition 3 = 0.86s, p_0,3_=0.023, p_1,2_<<0.001. Mann–Whitney U test).

**Figure 3.**
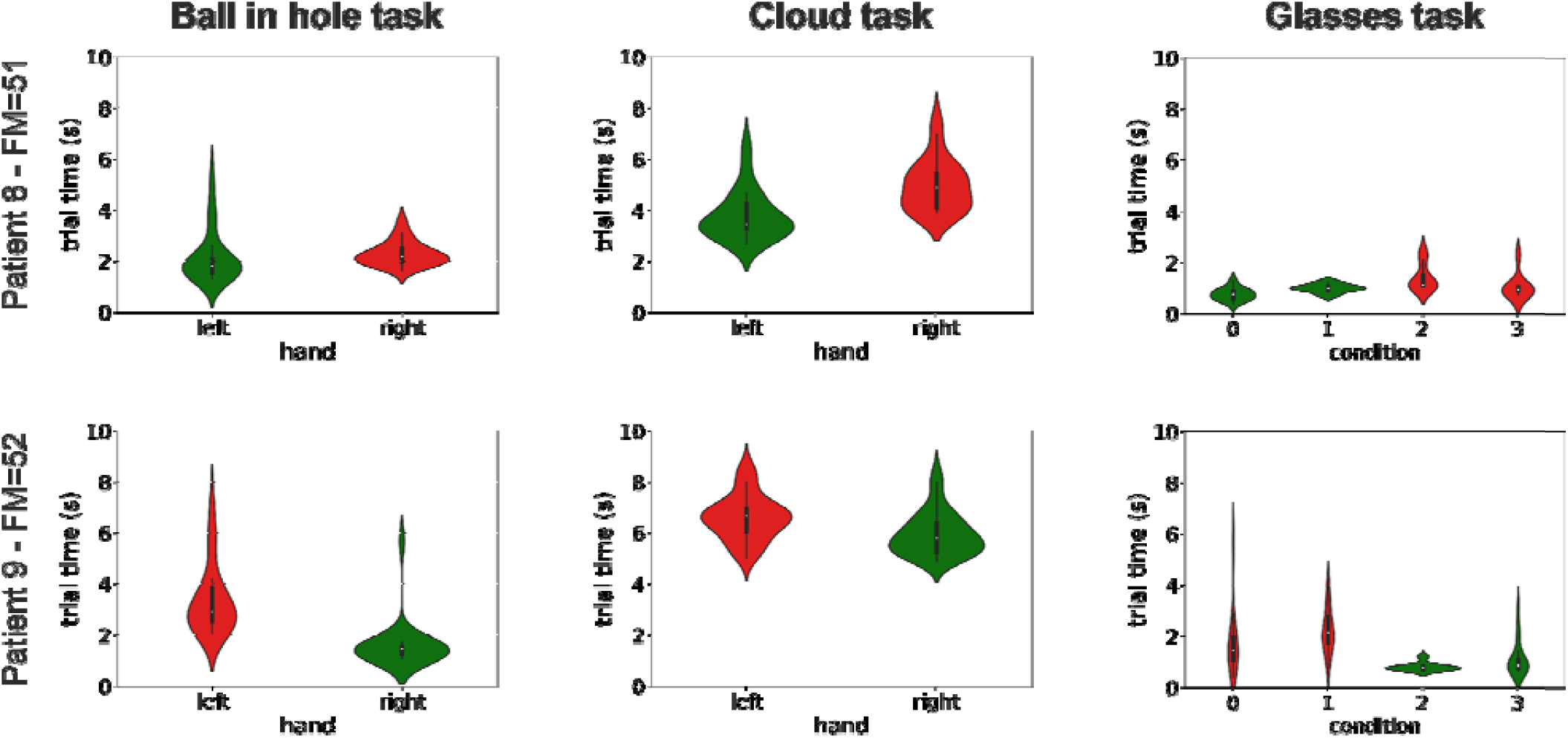
Distribution of completion times for three tasks and two patients. The violin plots show the distributions of the times taken to complete three of the tasks presently implemented in our system (three columns) for two patients. Patient 12 was a male in his 70’s with a right-side impairment, and patient 13 was female in her 60’s with a left-side impairment. Distributions are color coded differently for the healthy and impaired limb (green and red respectively). The label on the vertical axis shows the patient id and her/his Fugl-Meyer score. See Figure S1 in the supplementary information for similar plots for all other patients.

The results in Figure 3 suggest that completions times could be potentially used to assess the progress during the rehabilitation process. To explore this possibility, we performed a correlation analysis to investigate whether the presence of a negative correlation between the differences of the median completion times between the healthy and the impaired limb with the Fugl-Meyer score, one of the most widely used clinical assessment of upper limb motor recovery. The Fugl-Meyer score ranges from a minimum of 0 to a maximum of 66, with higher scores indicating less impairment. The results of our analysis are shown in Figure 4. Very interestingly, we found, even in our necessarily restricted pool of subjects, a significant negative correlation between differences in completion times and clinical scores for almost all tasks (ball in hole task: correlation=-0.66, p= 0.026; cloud task=-0.93, p<0.001; glasses task (pedestals 0 3): correlation=-0.82, p=0.003; glasses task (pedestals 1 2): correlation=-0.38, p=0.16; one-tailed Spearman’s rank-order test). The presence of a correlation between behavioral performances in our VR tasks and clinical scores suggests that the former, that are automatically computed by our system, can be conveniently used to measure progress during the rehabilitation process. This result is very promising and it suggests that, in addition to a higher degree of patients’ engagement, our system could also provide, in an automated manner, clinically meaningful indices of motor recovery to the rehabilitation therapists. Further studies in larger cohorts of patients are needed to fully validate this feature.

**Figure 4.**
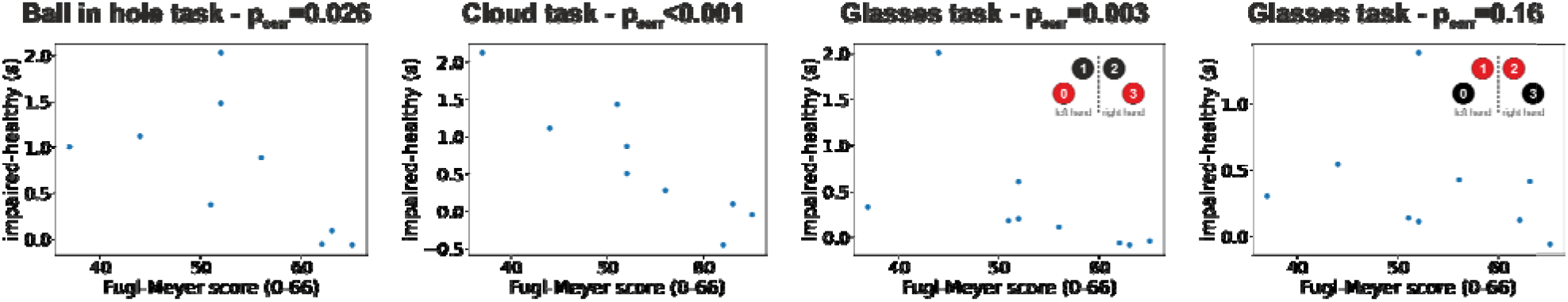
Correlation between behavioral results in our VR tasks and Fugl-Meyer clinical scores. The four scatterplots show the difference in completion times between the impaired and healthy limb for each patient and condition plotted against the Fugl-Meyer clinical assessment. Each panel shows results for one task and each dot represents data for one patients. The p-value of the correlation (Spearman’s rank-order correlation) between completion times and Fugl-Meyer scores is shown in the panels’ title.

Besides trial completion times, our system also saves the patients’ hand trajectories during task execution. In the Oculus Quest 2, hand positions are estimated visually. Their values exhibit thus larger errors compared to those recorded by more precise, but also more costly, motion capture systems. They can nonetheless be used to provide useful information to the therapist. For example, Figure 5 shows the hand trajectories recorded in a male patient in his 70’s during the performance of the Ball in hole (left panel) and Glass (right panel) tasks. As the figure shows, there are marked differences both in terms of movement span and smoothness between the trajectories of the impaired left arm and the healthy right arm. In addition to execution times, our system can thus also provide the therapist with relevant information concerning the trajectories of the patients’ arms that can be instrumental to assess the patient’s progress and inform the subsequent steps in the rehabilitation process.

**Figure 5.**
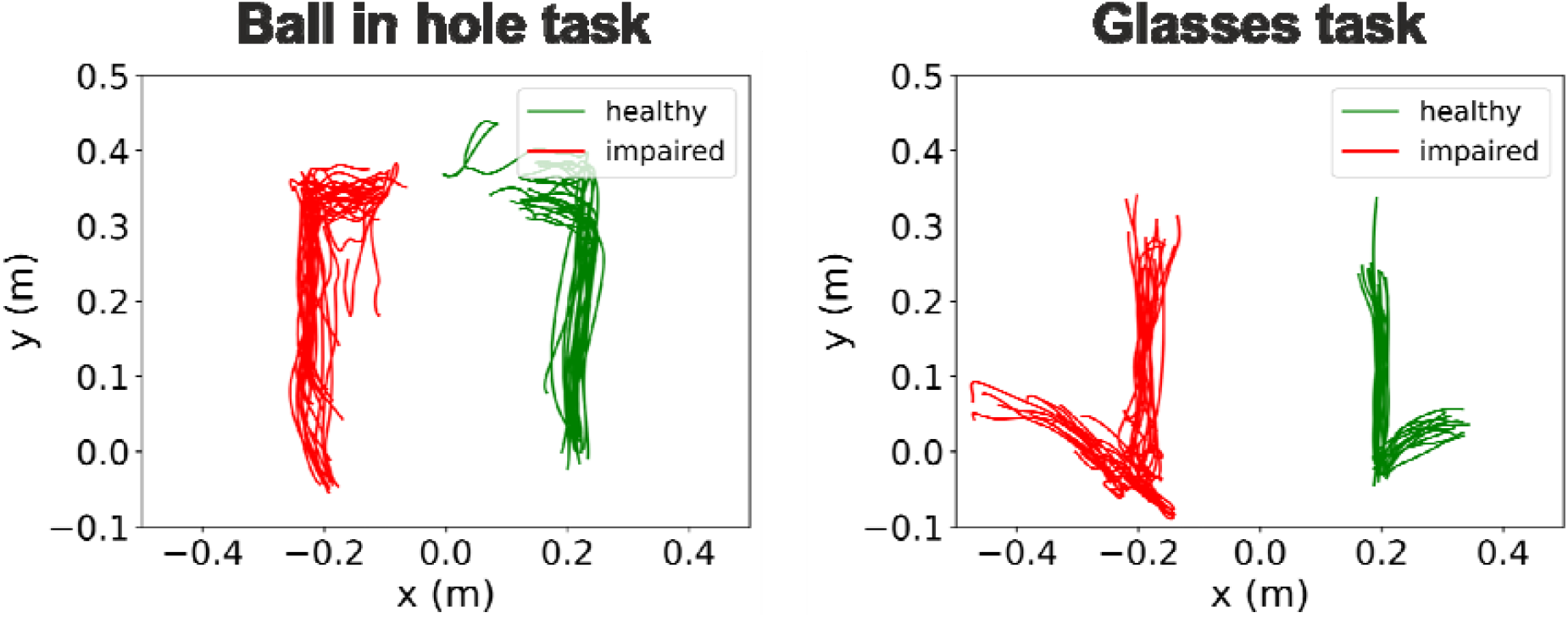
Example of hand trajectories recorded during task execution. The two panels show the hand trajectories of a male patient in his 70’s during the execution of the “ball in hole” (left panel) and “glasses” (right panel) tasks. This patient exhibited a left side impairment. The trajectories of the healthy and impaired hands are shown in green and red respectively.

## 4 Discussion

Here, we presented an innovative immersive virtual reality environment for upper limb rehabilitation (Figure 1) and we reported the results of a feasibility study in a group of 16 stroke patients. Almost all subjects gave the maximum rating to their experience (Figure 2A) and, in a standardized questionnaire (Gonzalez-Franco & Peck, 2018), they reported a high degree of ownership of the virtual hands and agency in the virtual environment (Figure 2B). Furthermore, we found that behavioral performances in our tasks, which can be automatically computed, correlate with the patients’ Fugl-Meyer clinical assessments (Figures 3 and 4). This suggests that, in the future, they could be effectively used as an automatically computed proxy of motor recovery. Notably, our system also stores the patients’ hand trajectories. Results in Figures 5 show that these data can be potentially used to provide valuable and quantitative information to the rehabilitation therapist to inform her/his decisions. Taken together, results presented here show that not only stroke patients enthusiastically accepted our VR system but also that it represents a promising and viable tool for upper limb motor rehabilitation. They motivate further studies to further validate its clinical efficacy.

A particularly interesting result of our experiments is that, for almost all of our tasks, the difference in task completion times between the impaired and healthy limb correlated with the Fugl-Meyer score, which is one of the most widely used clinical assessment of upper limb motor functions. This relationship suggests that differences in completion times can be used as a proxy of clinical scores, with two main advantages. First, while the computation of the Fugl-Meyer score requires a non-negligible amount of time and the involvement of specifically trained healthcare professionals, the differences in task completion times can be automatically computed by our system at the end, or even during, each training session. Second, given that they correlate with the Fugl-Meyer score, they can be used *within* a subject to monitor the efficacy of the rehabilitation process throughout its unfolding in time. In other words, our system can automatically provide ad interim clinically meaningful assessments of the progress of each patient, thus reducing the number of the more time- and resource-consuming clinical assessments.

It must be emphasized that the goal of our VR system is not to replace current rehabilitation therapies but rather to complement them and strengthen their efficacy (Fang et al., 2022) with a particular focus on two inter-related aspects: enhancing patients’ adherence and provide a viable option for telerehabilitation.

Rehabilitation therapies in post stroke patients often face adherence issues, in particular due to the need of exercises to be highly intensive and repetitive to effectively induce structural compensatory brain plasticity (Sampaio-Baptista et al., 2018). As such, they often become very tedious for the patients that end up complying only partially, or not at all, with what prescribed by the rehabilitation therapist (K. K. Miller et al., 2017). Gamification procedures have been shown to improve patients’ adherence to the rehabilitation schedules (da Silva Cameirão et al., 2011; Doumas et al., 2021). In this respect, more modern solutions based on immersive VR promise to deliver a more engaging experience to patients producing therefore higher adherence to the prescribed schedules. These solutions are presently gaining increasing traction (Crosbie et al., 2012; Mekbib et al., 2020; Ögün et al., 2019), as recent technical advancements have rendered virtual reality not only extremely realistic but also extremely cost-effective and ready for the consumer market. In addition, clinical studies have proven the effectiveness of these approaches (Laver et al., 2017). Our VR system is based on the Oculus Quest 2 state-of-the-art and off-the-shelf head-mounted display and, as such, it delivers an extremely realistic VR experience at a very accessible cost. In addition, it must be emphasized that, while the Oculus Quest 2 is presently our hardware of choice, the fact that we developed our VR-based rehabilitation system in the Unity development environment using, as much as possible, standard components, ensures that it can be ported to other HMDs with minimal efforts.

Telerehabilitation is a very interesting trend allowed by recent technological advancements. That is, moving part, or even most, of the rehabilitation procedures away from the hospitals, while maintaining medical supervision. Such process has benefits both for the patients and the hospitals. Throughout the rehabilitation period, stroke patients are required to move on a regular basis (i.e. 2-3 times a week) from their houses to a hospital or other healthcare institutions to perform motor rehabilitation sessions under the supervision of trained professionals. That is very taxing for stroke patients, who are, we must not forget, motor impaired, and it might produce additional non-adherence issues. Giving stroke patients an effective way to perform certified rehabilitation procedures at home would thus greatly contribute to increase their quality of life. This process would be also beneficial for the hospitals, as it would allow a better management of human and equipment resources, especially in view of handling potential future waves of Covid 19. With this respect, several features of our VR system were specifically implemented to support telerehabilitation scenarios. First, the control app (Figure 1B) communicates with the HMD via the internet. Thus, the computer running the app, controlled by the rehabilitation therapist, and the HMD, wore by the patient, can be in any place with the only requirement that they both have access to the internet. Second, the client app shows an exact replica of what is experienced by the patient in the VR environment. This provides the therapist with real-time information about task performance. Third, the therapist can vocally interact with the patients and set their schedule remotely and in real-time. Fourth, our VR system estimates and stores the patients’ hand trajectories during task performance. As shown in Figures 3-5, these data can potentially provide relevant information to the therapist and even provide quantitative and automatic assessments of the rehabilitation process. In summary, our VR system can not only greatly improve patients’ adherence to prescribed therapies but has been also specifically designed to support telerehabilitation scenarios.

As concerned about future progress of our VR rehabilitation system, the implementation of the mirror modality (see Methods section for details) can extend and increase the therapeutic applications in terms of patients’ subgroups and rehabilitative goals. The use of mirror therapy has shown clinical benefits in post-stroke patients in the improvement of upper limb motor function and impairment (Thieme et al., 2018), particularly for severely impaired ones (Colomer et al., 2016; Madhoun et al., 2020). This therapeutic intervention has proven to be instrumental also for pain reduction in patients affected by Complex Regional Pain Syndrome type 1 (Cacchio et al., 2009; Pervane Vural et al., 2016), a frequent and debilitating post-stroke condition that compromises rehabilitative outcomes. The use of immersive VR-based mirror therapy, which is characterized by a more intensive cognitive stimulation, may promote greater effects in these clinical conditions.

While we see many potential future developments for our VR-based rehabilitation system, a particularly interesting one is its combination with robotic platforms used in motor rehabilitation. These devices are becoming more widespread in the clinical practice and they provide a range of training conditions ranging from the passive resistance to the active assistance of single and multiple body segments during movements (Hesse et al., 2003; Iosa et al., 2012; Mehrholz et al., 2018). Robotic devices are presently routinely used in the clinical practice mainly for gait rehabilitation as they assist in supporting the patient’s bodily weight during training and help leg mobility (Calabrò et al., 2016). Furthermore, it has been shown that the combination of VR and gait-assisting devices enhances the activity of brain networks specifically involved in motor planning and learning (Calabrò et al., 2017). In the past, there attempts to combine arm exoskeletons and immersive virtual reality for the upper limb rehabilitation (Frisoli et al., 2009, 2007; Montagner et al., 2007). However, potentially due to the bulkiness and cost of exoskeletons, those attempts never translated to the clinical practice. In the past ten years robotic devices for upper limb rehabilitation have made consistent progress and they are presently not only used in the clinical practice, but their clinical efficacy has been proven by several studies (Mehrholz et al., 2018). There are thus presently exciting opportunities for combining them with immersive virtual reality and study whether this combination enhances, similar to the combination of gait training devices and VR, functional brain networks related to upper limb motor functions.

## Data Availability

All data produced in the present study are available upon reasonable request to the authors

## 6 Author Contributions

GF and NS collected the data. AB performed the clinical evaluations. AC and SS designed and supervised the study. AC designed and supervised the implementation of the VR system. AC and SS designed the VR tasks. AC, GF and NS analyzed the data. AC drafted the manuscript. All authors approved and finalized the manuscript.

## 8 Acknowledgments

We thank Luca Galofaro, Antonio Fogli and Nicolò Zironi for their contributions to software development and Chiara Paoluzzi for her help to collect patients’ data.

## Supplementary Material

### 1 Embodiment Questionnaire

The embodiment questionnaire was administered in Italian. It consisted of 6 questions derived from questions Q1, Q2, Q3, Q6, Q7, Q9 in (Gonzalez-Franco & Peck, 2018).

*“During the experiment there were moments in which…*

*Q1. I felt as the virtual hands were my hands”*

*Q2 I felt as the virtual hands were someone else’s”*

*Q3. I felt as I had more than two hands”*

*Q4. I felt as I could my virtual hands as it were my hands”*

*Q5. I felt as the movements of my virtual hands were caused by my hands”*

*Q6. I felt as the virtual hands were moving by themselves”*

### 2 Satisfaction Questionnaire

The satisfaction questionnaire was administered in Italian. It included 14 questions investigating pros and cons of the experience of the patients during the VR session. The numbering of the questions reflects the order in which they appeared in the questionnaire. Responses to question S1 are shown in Figure 1A in the main manuscript. Responses to other questions are shown in Figure S3 in the Supplementary Material.

English Version:

*Likert-scores questions (1-5):*

> *S1. Did you enjoy the training session?*
>
> *S2. Were suggestions during the exercise useful?*
>
> *S3. Was the training length appropriate?*
>
> *S4. Were the goals task easy to achieve?*
>
> *S5. Were the instructions easy to understand?*
>
> *S6. How many breaks did you take during the game session?*
>
> *S11. Are you physically tired?*
>
> *S12. Are you mentally tired?*
>
> *S13. How much fun did you have?*
>
> *S14. How satisfied with the training session are you?*

*Multiple-choices questions:*

> *S7. In which task did you perform best?*
>
> *S8. In which task did you perform worst?*
>
> *S9. Which task was most enjoyable?*
>
> *S10. Which task was the most boring?*

---

**Table S1.** Demographic and clinical characteristics of all patients in our study *(FMA-UE: Fugl Meyer Assessment – Upper Extremity)*.

| ID | SEX | AGE RANGE<br>(5 years) | STROKE TYPE | HEMIPARESIS SIDE | DAYS FROM STROKE EVENT | HOSPITALIZATION TYPE | FMA-UE score<br>(0-66) | VR SESSION (min) |
| --- | --- | --- | --- | --- | --- | --- | --- | --- |
| 1 | M | 61-65 | Hemorrhagic | Left | 382 | Day-Hospital | 43 | 45 |
| 2 | M | 56-60 | Ischemic | Right | 56 | Full | 34 | 35 |
| 3 | M | 56-60 | Ischemic | Right | 174 | Day-Hospital | 30 | 45 |
| 4 | M | 61-65 | Ischemic | Right | 66 | Full | 65 | 50 |
| 5 | F | 61-65 | Ischemic | Left | 106 | Day-Hospital | 30 | 45 |
| 6 | M | 56-60 | Ischemic | Left | 383 | Day-Hospital | 35 | 45 |
| 7 | F | 51-55 | Hemorrhagic | Right | 40 | Full | 60 | 45 |
| 8 | M | 76-80 | Ischemic | Right | 32 | Full | 51 | 40 |
| 9 | F | 61-65 | Ischemic | Right | 39 | Full | 52 | 60 |
| 10 | M | 76-80 | Ischemic | Left | 149 | Full | 37 | 60 |
| 11 | M | 61-65 | Ischemic | Right | 24 | Full | 65 | 60 |
| 12 | M | 66-70 | Ischemic | Left | 75 | Full | 44 | 45 |
| 13 | M | 61-65 | Hemorrhagic | Left | 92 | Full | 63 | 60 |
| 14 | M | 76-80 | Hemorrhagic | Left | 92 | Full | 52 | 45 |
| 15 | M | 41-45 | Hemorrhagic | Left | 196 | Full | 62 | 60 |
| 16 | F | 51-55 | Ischemic | Right | 4216 | Day-Hospital | 56 | 60 |

**Figure S1.**
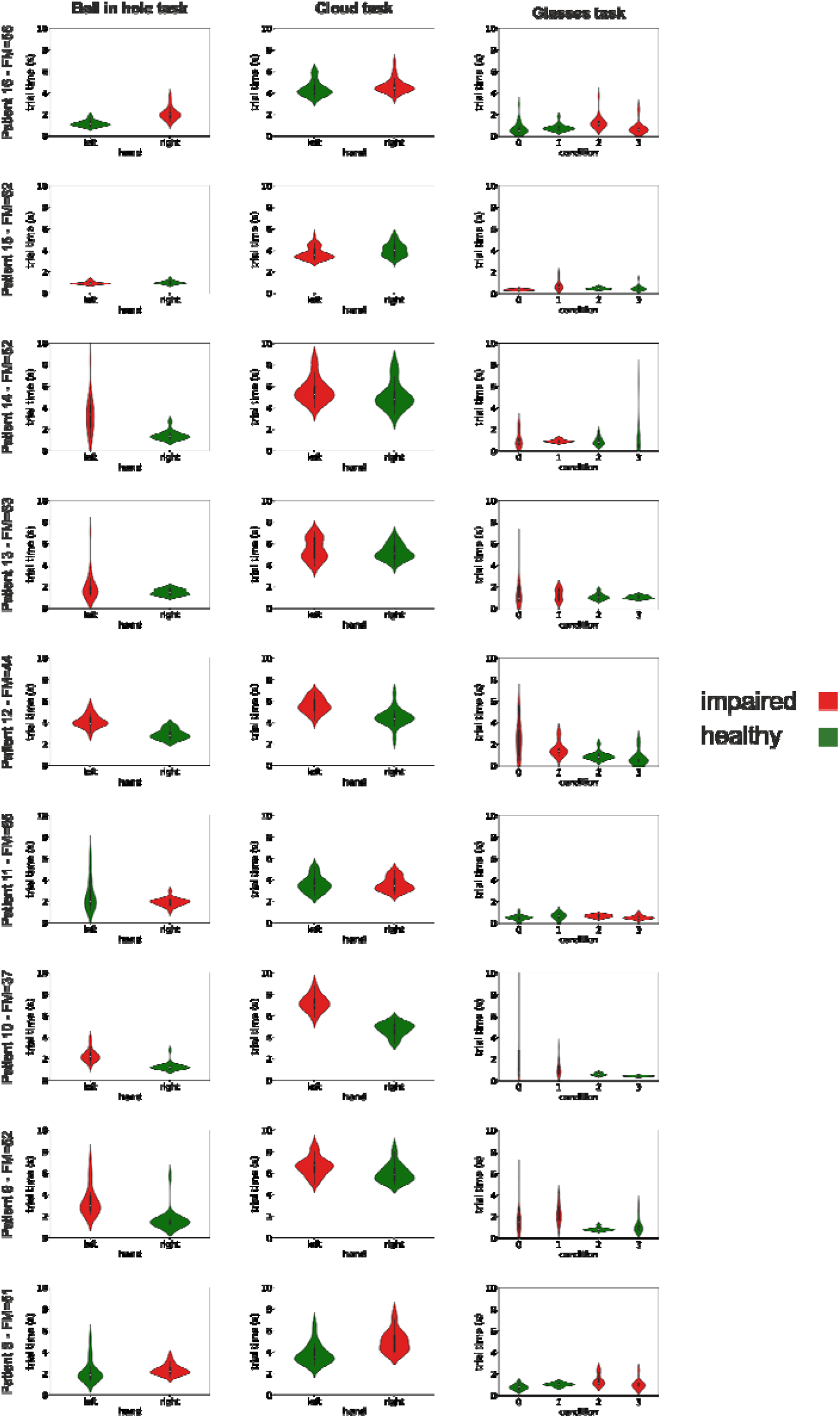
Distribution of completion times for three tasks and all patients for which completion times were recorded (9 out of 16 patients). Labels and conventions are as in Figure 3.

**Figure S2.**
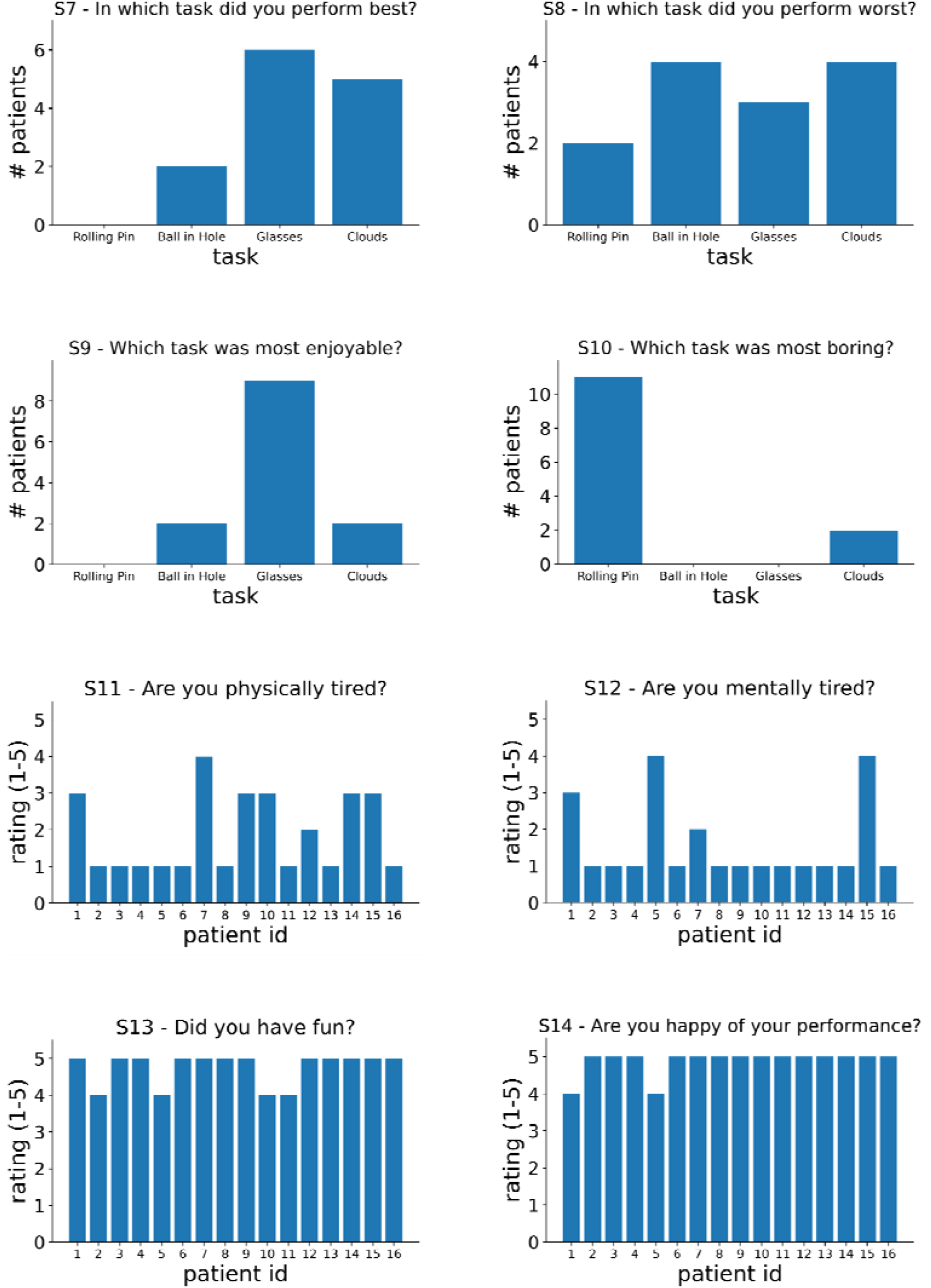
Patients’ responses to questions in the satisfaction questionnaire.

